# Rumination Induction Task in fMRI: Test-Retest Reliability in Youth and Potential Mechanisms of Change with Intervention

**DOI:** 10.1101/2023.10.09.23296759

**Authors:** Mindy Westlund Schreiner, Raina H. Miller, Anna M. Jacobsen, Sheila E. Crowell, Erin A. Kaufman, Brian Farstead, Daniel A. Feldman, Leah Thomas, Katie L. Bessette, Robert C. Welsh, Edward R. Watkins, Scott A. Langenecker

**Affiliations:** Department of Psychiatry, Huntsman Mental Health Institute, University of Utah; Department of Psychology, University of Utah; Department of Biomedical Engineering, University of Utah; Department of Psychology, University of Oregon; Department of Psychiatry & Behavioral Health, The Ohio State University; Department of Psychiatry & Biobehavioral Sciences, University of California Los Angeles; Department of Experimental & Applied Clinical Psychology, University of Exeter

## Abstract

**Background:** Rumination is a transdiagnostic problem that is common in major depressive disorder (MDD). Rumination Focused Cognitive Behavioral Therapy (RF-CBT) explicitly targets the ruminative habit. This study examined changes in brain activation during a rumination induction task in adolescents with remitted MDD following RF-CBT. We also evaluated the reliability of the rumination task among adolescents who received treatment as usual (TAU).

**Method:** Fifty-five adolescents ages 14-17 completed a self-relevant rumination induction fMRI task and were then randomized to either RF-CBT (n = 30) or TAU (n = 25). Participants completed the task a second time either following 10-14 sessions of RF-CBT or the equivalent time delay for the TAU group. We assessed activation change in the RF-CBT group using paired-samples t-tests and reliability by calculating intraclass correlation coefficients (ICCs) of five rumination-related ROIs during each of three blocks for the TAU and RF-CBT groups separately (Rumination Instruction, Rumination Prompt, and Distraction).

**Results:** Following treatment, participants in the RF-CBT group demonstrated an increase in activation of the left precuneus during Rumination Instruction and the left angular and superior temporal gyri during Rumination Prompt (*p* < .01). The TAU group demonstrated fair to excellent reliability (*M* = .52, range = .27-.86) across most ROIs and task blocks. In contrast, the RF-CBT group demonstrated poor reliability across most ROIs and task blocks (*M* = .21, range = -.19-.69).

**Conclusion:** RF-CBT appears to lead to rumination-related brain change. We demonstrated that the rumination induction task has fair to excellent reliability among individuals who do not receive an intervention that explicitly targets the ruminative habit, whereas reliability of this task is largely poor in the context of RF-CBT. This has meaningful implications in longitudinal and intervention studies, particularly when conceptualizing it as an important target for intervention. It also suggests one of many possible mechanisms for why fMRI test-retest reliability can be low that appears unrelated to the methodology itself.

## Rumination Induction Task in fMRI: Effects of Rumination Focused Cognitive Behavioral Therapy and Test-Retest Reliability in Youth

Rumination, or repetitive negative thinking, has been of increasing interest among psychopathology researchers. Repetitive negative thinking may be a transdiagnostic marker for psychopathology risk, especially for mood disorders. Continued rumination after recovery from a major depressive episode is a significant risk factor for depression recurrence (Figueroa et al., 2019). Thus, rumination may be stable in the context of both active and remitted psychopathology. As such, targeting rumination may be fruitful for enacting meaningful change *and* reducing future psychopathology risk.

Encouragingly, treatments that specifically target rumination reduction have illustrated promising short- and long-term benefits. For example, there was a 62% remission rate after just 12 sessions of Rumination Focused Cognitive Behavioral Therapy (RF-CBT) in a phase II randomized controlled trial (Watkins, 2018). One RF-CBT randomized controlled trial in adolescents with pre- and post-intervention fMRI scans found youth reported reduced depression and rumination after 8 sessions (Jacobs et al., 2016). This study also showed a significant reduction in connectivity between the left posterior cingulate cortex (PCC) and the right inferior frontal gyrus (IFG) and bilateral inferior temporal gyri (ITG) during resting-state fMRI (Jacobs et al., 2016). Changes in self-reported depression and rumination were correlated with degree of change in connectivity, indicating that differences in default mode network (DMN) activity are involved in rumination and depression. This connectivity and rumination correlation has recently been replicated in an independent sample by the same group (Langenecker et al., in press), suggesting a common neural correlate for reductions in rumination.

While the utility of resting-state fMRI is well established, fMRI tasks have been reported to have poor reliability upon retest. In particular, a meta-analysis by Elliott and colleagues demonstrated poor reliability across a variety of tasks, including picture naming, emotion identification and matching, button pressing, working memory, episodic memory, and reward (Elliott et al., 2020). Further, when examining tasks used in large imaging cohort studies (Dunedin Study and Human Connectome Project), aggregated reliability was again typically poor. One challenge to take away from such work is that aggregated reliability values pertain to only a small subset of tasks available for use in fMRI literature. There are many potential catalysts for poor test reliability in fMRI tasks. This includes time pressures in the scanner (too few events), movement within the scanner (measuring different neural systems), and physiological sources of variability (heart rate). Yet, there are other sources of potential disruption to reliability. In the context of intervention work, we would expect and intend to see pre- to post-treatment changes that might drive down reliability. Here, lower reliability in a randomized active treatment arm acts as a therapeutic intervention manipulation check, rather than an issue inherent to the fMRI measure being used. Further, task reliability, or lack thereof, may highlight mechanisms of action (and brain locations) underlying therapeutic success. It is possible to use differential reliability estimates in a treatment condition compared to a time matched comparison group to dissociate 1) Test-retest reliability of relevant neural circuits, and 2) Pathways by which treatment induces change. This highlights the potentially unique importance of understanding the test-retest reliability of an fMRI task—without good reliability it cannot be considered an important target for intervention.

In contrast to the summary by Elliott and colleagues (2020), studies have highlighted that fMRI task reliability may be dependent on brain regions of interest and the specific tasks that are used to elicit activation within those areas. Certain brain regions may be more likely to demonstrate task reliability over time. For example, researchers have consistently found good to excellent reliability in the occipital regions across fMRI tasks (Herting et al., 2018) while a recent meta-analysis synthesized fMRI findings from brain regions implicated in rumination (Zhou et al., 2020) and found relevant brain regions demonstrated fair to high reliability. Further, one longitudinal study found that both adolescents and adults demonstrated fair to good reliability in bilateral precuneus, left inferior and superior parietal cortices, and right angular gyrus in a rule-switch task (Koolschijn et al., 2011). The middle temporal gyrus has also shown high test-retest reliability in an emotion regulation task (Berboth et al., 2021). Other research indicates that attentional switching ability, a cognitive process implicated in rumination, has moderate to high test-retest reliability and is fairly stable (Koster et al., 2013).

Taken together, findings indicate that brain regions and cognitive processes involved in rumination have moderate to high test-retest reliability. Thus, a rumination induction fMRI task may also have adequate test-retest reliability, particularly in the absence of intervention. Indeed, one study found that a rumination induction task yielded reproducible findings across different scanners (Chen et al., 2020). Further, Bessette and colleagues demonstrated medium to large correlations of brain activation across blocks within the rumination induction task among adolescents (Bessette et al., 2020). However, assessment of test-retest reliability of rumination induction tasks is still needed. Existing neurobiological research on rumination has implicated brain regions including the precuneus, anterior cingulate cortex (ACC), paracingulate, inferior frontal gyrus (IFG), angular gyrus, and superior temporal gyrus (STG). These regions are involved in self-referential processing, memory retrieval, and emotional processing, which all play a role in the ruminative habit. While work has demonstrated significant changes in resting- state functional connectivity following an intervention that targets rumination, little work has evaluated neurobiological changes in response to fMRI tasks where rumination is *explicitly* induced.

The present study sought to 1) evaluate change in neurobiological responses to a common rumination induction task among adolescents who received RF-CBT, and 2) examine the reliability of this rumination induction task in the group of adolescents who were not randomized to the RF-CBT arm of this trial. These data are derived from a larger study investigating the utility of RF-CBT for reducing rumination in a sample of adolescents with remitted MDD and high levels of self-reported rumination (to reduce likelihood of depression recurrence; Langenecker et al., in press). Based on a prior meta-analysis of brain regions associated with rumination (Zhou et al., 2020), we specifically examined changes and reliability in five regions of interest (ROIs): 1) left precuneus, 2) left STG, 3) left ACC and paracingulate, 4) left angular gyrus, and 5) left IFG. In this randomized control trial, participants completed MRI scanning and were either randomized to 10-14 sessions of RF-CBT or to Treatment as Usual (TAU).

Participants then completed a second, post-intervention MRI. We anticipated that the RF-CBT group would demonstrate significant change in activation of these five ROIs during the rumination induction task, and thus possibly poor reliability between Time 1 and Time 2 fMRI. Conversely, we expected brain activation during the rumination induction task would show high reliability between Time 1 and Time 2 in the TAU group.

## Method

### Participants

Data were collected as part of a larger clinical trial (Langenecker et al., in press; NCT03859297). We recruited adolescents with remitted major depressive disorder from Salt Lake City and surrounding areas through Facebook, radio, and community advertisements. Eligible participants were 14-17 years of age with a history or major depressive disorder (currently either in full or partial remission). Exclusion criteria consisted of lifetime history of mania or psychosis, suicide attempt in the past 6 months, current active suicidal plan or intent, a score above 45 on the Children’s Depression Rating Scale-Revised (CDRS-R; Poznanski & Mokros, 1996), developmental disorders such as autism spectrum disorder and pervasive developmental delay, and MRI contraindications. As this was a clinical trial, exclusion criteria also included any CBT in the past six months and changes in psychiatric medications within 6 weeks prior to enrollment (with no dosage change in the prior 4 weeks). All participants were encouraged to maintain their treatment as usual for the duration of the study.

### Measures

#### Clinical Assessment

Participants and at least one parent or guardian completed a clinical assessment with a trained clinician. Assessments included the Kiddie-Schedule for Affective Disorders and Schizophrenia-Present and Lifetime Version (KSADS-PL; Kaufman et al., 1997) to determine current and past psychopathology, and the Children’s Depression Rating Scale-Revised (CDRS-R; Poznanski & Mokros, 1996) to determine current depressive symptoms.

#### Neuroimaging

Participants completed MRI scanning at the Imaging and Neurosciences Center at the University of Utah, where we acquired simultaneous-multi-slice data (Feinberg & Setsompop, 2013). The scanning protocol included a high-resolution T1 structural image, which we used for registration of the task data. The T1 was acquired with the following parameters: field of view (FOV) = 256mm, TE = 2500ms, TR = 2500ms, flip angle = 8 degrees, GeneRalized Autocalibrating Partial Parallel Acquisition (GRAPPA) acceleration factor = 2 (Griswold et al., 2002), multiband factor = 4, and voxel size = 0.8mm isotropic.

Participants completed the Rumination Induction Task, which is a 9-minute block design completed in a single run with rumination and distraction conditions. We recently published some of these data, which examined the association between brain activation and non-suicidal self-injury (Westlund Schreiner et al., 2023). Our group has also used this task successfully with other adolescent samples (Bessette et al., 2020; Burkhouse et al., 2017). Before entering the scanner, participants are asked to generate four events from their own lives: a sad event, a frustrating event, a failure event, and a hurtful event. As these events are designed to elicit rumination, we had participants rate each event on a 9-point scale regarding how upset the event made them, both at the time it was happening, and at the current moment (1 = *Did not feel sad* and 9 = *Felt very sad).* When past or current ratings were rated as less than five, participants were encouraged to choose different memories to ensure sufficient potency to elicit rumination.

When in the scanner, participants are asked to bring one of the four memories to their mind in detail over a 25 second block (Rumination Instruction). They are then directed to reflect on this past event using prompts over the course of 30 seconds (e.g., “Think about what your feelings mean”, “Think about why you reacted the way you did”, etc.; Rumination Prompt).

Following the Rumination Prompt blocks, they are asked “How much are you focused on your feelings right now?” and “How sad do you feel right now?” using a scale of 1-5 (1 = *not focused on feelings/sad*, 5 = *very focused on feelings/sad*). This was followed by a 10-15 second block during which participants are instructed to “Rest” and a 30 second distraction prompt (e.g., “Think of a row of shampoo bottles”; Distraction). Participants completed ratings about how focused they were on their feelings and how sad they felt following each Distraction block.

Participants completed a total of 4 repetitions of these conditions (one for each event) during the task (see Figure 1).

**Figure 1.**
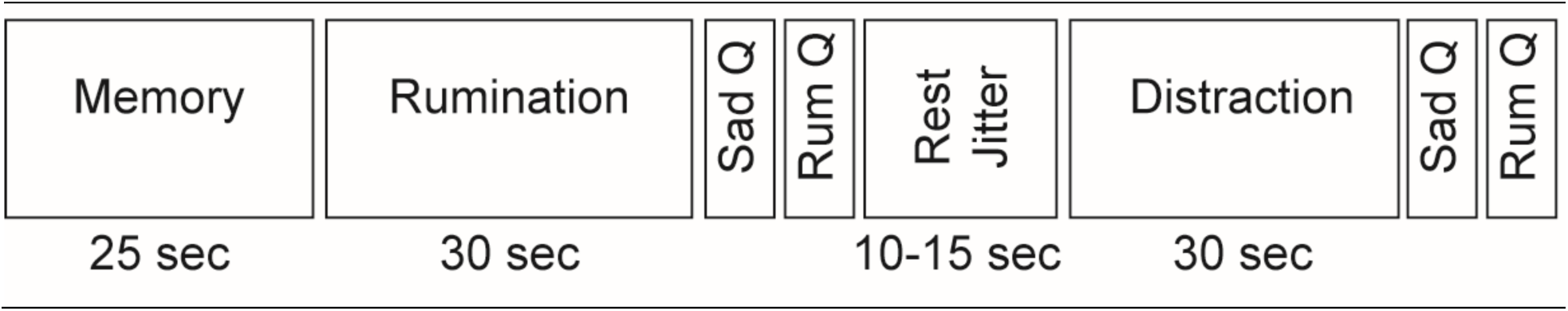
Rumination Induction Task. Each full sequence completed 4 times.

### Procedure

This project was approved by the Institutional Review Board at the University of Utah. Persons interested in enrollment first completed a brief phone screen to determine eligibility. If eligible, participants were scheduled for their initial visit where they completed informed written consent and assent. Following consent/assent, youth and their parent/guardian completed self-report measures and a clinical assessment with a trained independent evaluator to gather information regarding their current and past mental health symptoms. Youth participants completed the MRI scan at a separate visit, during which they completed the rumination induction task. As part of the randomized clinical trial, youth were randomized to either 10-12 sessions of RF-CBT or TAU. After completing RF-CBT (or after approximately 3-4 months for the TAU group), adolescents completed a second clinical assessment and MRI visit. Participants were compensated for their time for all non-treatment related visits.

### Analyses

#### Neuroimaging Preprocessing and Analysis

We used tools from Anima (Voss et al., 2006) and Statistical Parametric Mapping 12 (SPM12; https://www.fil.ion.ucl.ac.uk/spm/doc/) for preprocessing and first- and second-level analyses. Consistent with recent work (Westlund Schreiner et al., 2023) we completed preprocessing including echo-planar imaging (EPI) distortion correction, discarding the first 10 volumes of functional data, time-series realignment, high-resolution T1 co-registration, high-resolution T1 tissue segmentation, normalization of high-resolution and functional images to standard space (MNI), and 5mm Gaussian smoothing. We used SPM12 to complete first-level whole-brain activation analyses with the rumination induction data. This included modeling key blocks of interest (Rumination Instruction, Rumination Prompt, and Distraction) and fixation, as well as including six motion parameters as covariates.

For second-level analyses, we created the five ROIs using the coordinates independently derived and published by Zhou and colleagues (Zhou et al., 2020; see Table 1 for ROIs and MNI coordinates). We specified a radius of 10mm for each region except for the angular gyrus, where we used a radius of 5mm. We used MarsBaR within SPM12 to create the ROIs and extract values for the Rumination Instruction, Rumination Prompt, and Distraction blocks for all participants’ Time 1 and Time 2 data (Brett et al., 2010).

**Table 1.**
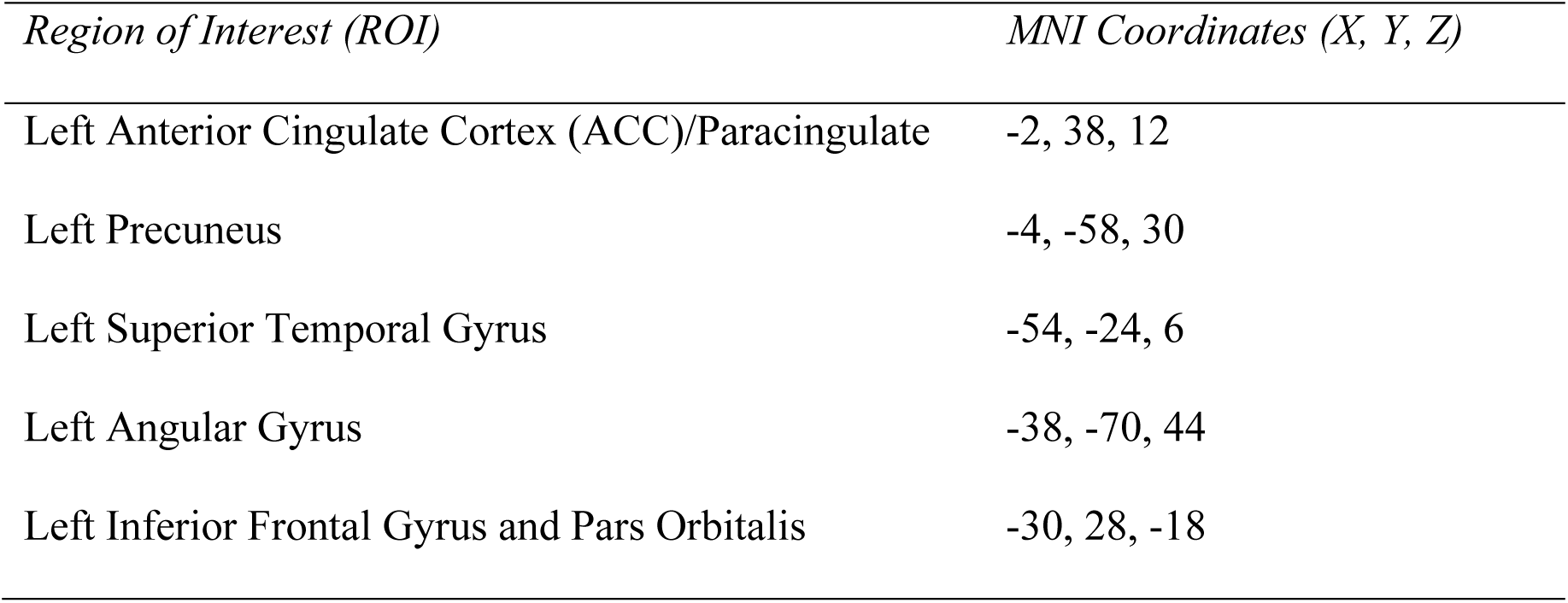
Regions of Interest and Coordinates.

| *Region of Interest (ROI)* | *MNI Coordinates (X, Y, Z)* |
| --- | --- |
| Left Anterior Cingulate Cortex (ACC)/Paracingulate | -2, 38, 12 |
| Left Precuneus | -4, -58, 30 |
| Left Superior Temporal Gyrus | -54, -24, 6 |
| Left Angular Gyrus | -38, -70, 44 |
| Left Inferior Frontal Gyrus and Pars Orbitalis | -30, 28, -18 |

#### Demographics and Clinical Characteristics

After we extracted the values from our five regions of interest, we combined these data with our demographic and clinical data. We used R to complete analyses, which include descriptive statistics for demographic and clinical data (R Core Team, 2022).

#### Treatment Change and Reliability

To evaluate whether there were significant changes in brain activation during the rumination induction task following RF-CBT, we conducted paired-samples t-tests in the RF- CBT group. We used the *irr* package in R to calculate intraclass correlation coefficients between the Time 1 and Time 2 data for the rumination induction task (Gamer et al., 2019; R Core Team, 2022). We calculated ICCs under the assumption of a two-way mixed effects model (ICC(3,1); Shrout & Fleiss, 1979) using consistency in measurement as opposed to agreement in absolute values (ICC(C,1); McGraw & Wong, 1996). We interpreted the ICC values based on previously established guidance in which ICC < .4 = Poor, .4 < ICC < .59 = Fair, .6 < ICC < .74 = Good, .75 > ICC > 1 = Excellent (Cicchetti, 1994; Cicchetti & Sparrow, 1981).

## Results

### Demographics and Clinical Characteristics

Fifty-five participants completed the rumination induction task at both time points. Thirty were randomized the RF-CBT condition and 25 were randomized to TAU. Table 2 provides additional information about the sample. Further information about the larger clinical trial can be found in Langenecker et al. (in press). There was no significant difference in number of days between scans for TAU (*M* = 188, *SD* = 43.3) and RF-CBT (*M* = 126, *SD* = 28.2), *t*(39.87) = - 0.70, *p* = .49.

**Table 2.** Demographic Characteristics.

|  | **TAU**<br>**(N=25)** | **RF-CBT**<br>**(N=30)** | **Overall**<br>**(N=55)** |
| --- | --- | --- | --- |
| **Mean Age (SD)** | 16.1 (0.971) | 15.8 (0.986) | 16.0 (0.981) |
| **Sex** |  |  |  |
| Male | 8 (32.0%) | 8 (26.7%) | 16 (29.1%) |
| Female | 17 (68.0%) | 22 (73.3%) | 39 (70.9%) |
| **Gender** |  |  |  |
| Female (Cisgender) | 8 (32.0%) | 14 (46.7%) | 22 (40.0%) |
| Male (Cisgender) | 2 (8.0%) | 8 (26.7%) | 10 (18.2%) |
| Transgender Male | 1 (4.0%) | 0 (0%) | 1 (1.8%) |
| Transgender Female | 0 (0%) | 1 (3.3%) | 1 (1.8%) |
| Non-Binary | 2 (8.0%) | 2 (6.7%) | 4 (7.3%) |
| Other | 2 (8.0%) | 1 (3.3%) | 3 (5.5%) |
| Missing | 10 (40.0%) | 4 (13.3%) | 14 (25.5%) |
| **Sexuality** |  |  |  |
| Straight (Heterosexual) | 7 (28.0%) | 17 (56.7%) | 24 (43.6%) |
| Gay | 0 (0%) | 1 (3.3%) | 1 (1.8%) |
| Lesbian | 1 (4.0%) | 1 (3.3%) | 2 (3.6%) |
| Queer | 2 (8.0%) | 1 (3.3%) | 3 (5.5%) |
| Pansexual | 3 (12.0%) | 4 (13.3%) | 7 (12.7%) |
| Asexual | 1 (4.0%) | 0 (0%) | 1 (1.8%) |
| Bisexual | 1 (4.0%) | 2 (6.7%) | 3 (5.5%) |
| Other | 0 (0%) | 1 (3.3%) | 1 (1.8%) |
| Missing | 10 (40.0%) | 3 (10.0%) | 13 (23.6%) |
| **Race** |  |  |  |
| White | 23 (92.0%) | 26 (86.7%) | 49 (89.1%) |
| Black/African American | 0 (0%) | 0 (0%) | 0 (0%) |
| Asian | 0 (0%) | 2 (6.7%) | 2 (3.6%) |
| Native Hawaiian or Other Pacific Islander | 0 (0%) | 0 (0%) | 0 (0%) |
| American Indian or Alaska Native | 0 (0%) | 1 (3.3%) | 1 (1.8%) |
| Other | 0 (0%) | 1 (3.3%) | 1 (1.8%) |
| Missing | 2 (8.0%) | 0 (0%) | 2 (3.6%) |
| **Ethnicity** |  |  |  |
| Hispanic or Latin(x) | 3 (12.0%) | 5 (16.7%) | 8 (14.5%) |
| Not Hispanic or Latin(x) | 20 (80.0%) | 25 (83.3%) | 45 (81.8%) |
| Missing | 2 (8.0%) | 0 (0%) | 2 (3.6%) |

### Brain Activation Change during Rumination Induction Following RF-CBT

The RF-CBT group demonstrated significant increases in activation following treatment across multiple ROIs. During both the Rumination Instruction and Rumination Prompt blocks, post-treatment activation values were higher in the left angular gyrus, precuneus, and STG (see Table 3). When using a Bonferroni-corrected p-value threshold of .01 (accounting for five ROIs), activation differences for the left precuneus during the Rumination Instruction blocks and the left angular gyrus and superior temporal gyrus during the Rumination Prompt blocks remained significant. There were no significant changes over time for activation during the Distraction blocks.

**Table 3.**
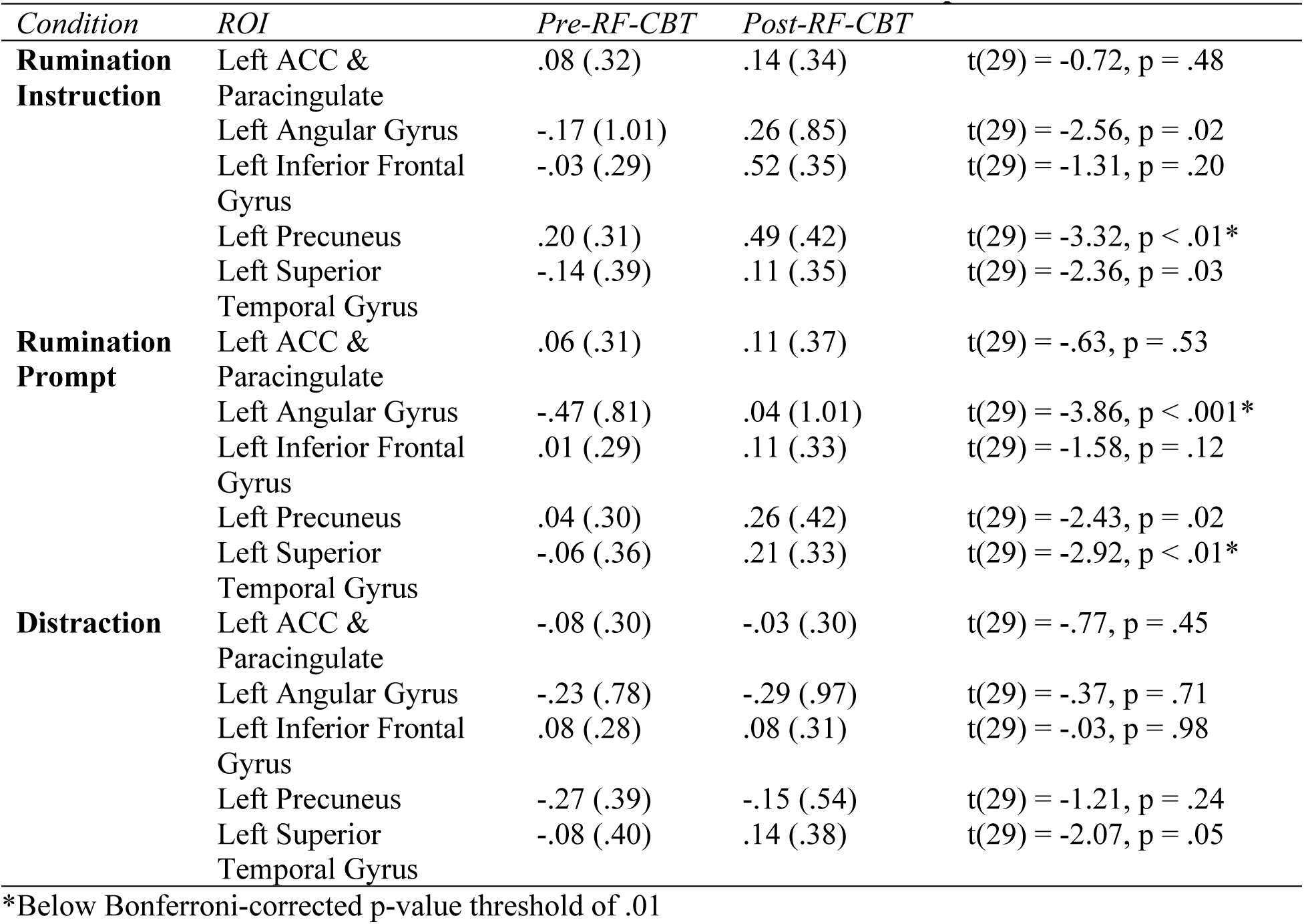
Pre- versus Post-Treatment Activation in RF-CBT Group.

### Task Reliability

Task reliability intraclass correlation coefficients are reported in Table 4. Consistent with our expectations the TAU group demonstrated fair to excellent test-retest reliability across nearly all ROIs and task blocks. The Rumination Instruction blocks demonstrated the highest reliability. During Rumination Instruction, activation of the left angular gyrus demonstrated excellent reliability, followed by the left precuneus which demonstrated good reliability. The remaining three ROIs demonstrated fair reliability. For the Rumination Prompt and Distraction blocks, the left angular gyrus demonstrated good reliability and the left precuneus demonstrated fair reliability. The left ACC and paracingulate demonstrated fair reliability during the Rumination Prompt and good reliability during Distraction. The left STG demonstrated fair reliability during the Distraction blocks and poor reliability during the Rumination Prompt. The left IFG demonstrated poor reliability for both Rumination Prompt and distraction blocks.

**Table 4.**
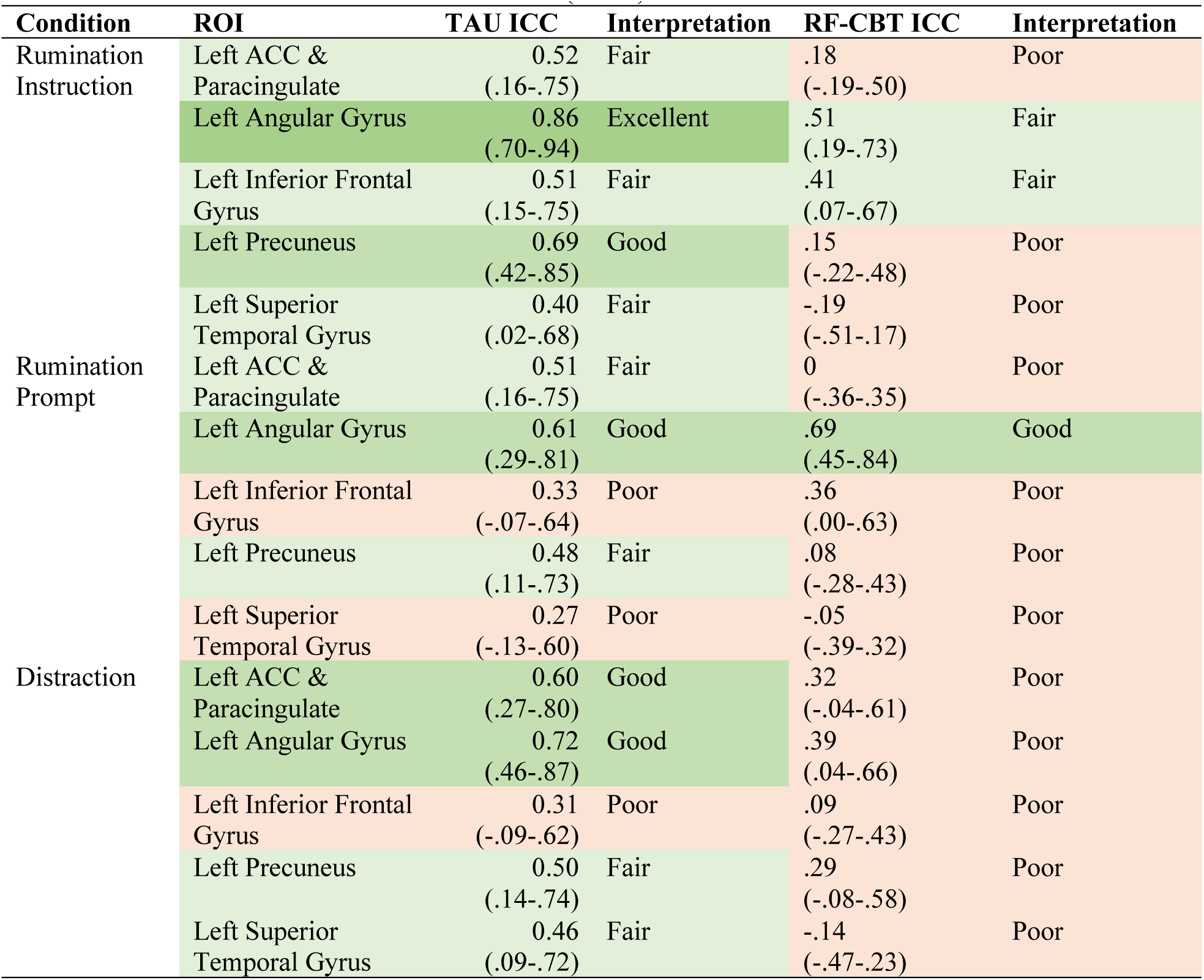
Intraclass Correlation Coefficients (ICCs) for Rumination Induction Task.

Reduced reliability, potentially associated with intentional treatment change, was observed in the RF-CBT group. The poor test-retest reliability in RF-CBT was observed for most ROIs and task blocks. Indeed, only the left angular gyrus and IFG showed fair reliability during Rumination Instruction and the left angular gyrus showed good reliability during Rumination Prompt blocks. This is consistent with our hypotheses as we anticipated that specifically targeting the ruminative habit would lead to changes in rumination related neural activation, AND reductions in reliability for these key regions.

## Discussion

The present study provides three encouraging and potentially provocative results. First, we provide evidence that there is acceptable fMRI reliability of the rumination induction task in key regions associated with rumination derived from prior empirical studies. Second, strong reliability in these rumination-related regions is specifically related to youth who did *not* receive a treatment that was intended to *change* the rumination habit. In contrast, reliability was poor in those who specifically *did* receive a treatment developed to change the rumination habit and related brain activation. Third, activation in rumination-associated brain regions significantly *increased* during rumination following RF-CBT.

The present study found that youth with remitted MDD (and elevated rumination) who completed RF-CBT showed increased activation of the angular gyrus, precuneus, and STG during the rumination induction task relative to pre-treatment. As brain activation is a relative measure in the context of fMRI tasks, this makes the present findings difficult to interpret. While an *increase* in activation of rumination-related brain regions would seem antithetical to RF-CBT, it is possible that this is instead a reflection of a greater contrast between blocks in which participants are ruminating and blocks during which they are not given explicit instruction (i.e., Rest blocks). In this case, results would suggest that following RF-CBT, participants were able to better disengage from rumination during non-rumination task blocks. A body of literature suggests that rumination persistence may relate to diminished ability to switch or inhibit mental sets, which could be related to difficulty disengaging relevant neural circuitry (Hilt et al., 2014; Whitmer & Banich, 2007). The increase and positive activation in rumination relevant regions during rumination blocks following RF-CBT suggests that RF-CBT may be facilitating greater dynamic range and flexibility in these rumination-related nodes.

Given recent concerns regarding the reliability of fMRI tasks, we found it necessary and valuable to evaluate the reliability of the rumination induction task. This is especially critical in the context of longitudinal and interventional studies as it facilitates more confident interpretations when attributing task activation changes to either treatment or a particular characteristic, rather than fMRI noise. Using the TAU group of this clinical trial as a benchmark, results indicate fair reliability of the rumination induction task, with certain brain regions showing higher test-retest reliability than others. In particular, the angular gyrus and precuneus had good to excellent reliability during the Rumination Instruction blocks. Since RF-CBT specifically targets rumination, we also examined whether the reliability of the rumination induction task was poor within the RF-CBT group.

There are also several reflections and limitations relevant to this work. First, it is notable that this task is a block design, which tends to have stronger test-retest reliability than event-related designs. Second, the task was developed after a strong base of clinical studies demonstrating psychological and clinical validity (Cooney et al., 2010; Johnson et al., 2006; Lyubomirsky & Nolen-Hoeksema, 1993). Thus, conversion to fMRI conditions was predicated on knowledge that the task was already valid and reliable. Third, this is a homogeneous sample of teens with higher levels of rumination and history of depression. While such homogeneity might be expected to reduce range and variability, constraining reliability, that was not observed here. Fourth, this is a relatively modest sample size for test-retest reliability. It is, however, relatively large compared to similar intervention studies in youth with neuroimaging.

In conclusion, we found strong reliability for the brain regions where we expected stable activation and poor test-retest reliability across ROIs and task blocks where we intended to elicit poor stability, in alignment with our hypotheses. Therefore, RF-CBT appears to specifically change rumination related circuitry during state-induced rumination, offering exciting avenues for future work that integrates neuroimaging and intervention. It also appears to be the case that the rumination induction task is likely measuring aspects of both state (amenable to treatment-related change) and trait (not amenable to treatment-related change) rumination. With our team’s ongoing collection of a larger sample of youth who are receiving either RF-CBT or Relaxation Therapy, we can further explore this relation (NCT03859297; R33MH116080). Other longitudinal studies of youth with multiple time points may further support the use of this task as a reliable measure of rumination.

## Acknowledgments

The authors would like to thank the participants and their families for contributing their time toward this project. We are also thankful for staff, volunteers, and students within the Multifaceted Explorations of the Neurobiology of Depressive Disorders (MEND2) Lab who have made this work possible.

## Ethics Statement

This study was approved by the Institutional Review Board at the University of Utah

## Data Availability Statement

These data are from a NIMH-funded study and will be available through the NIMH Data Archive. Data are also available upon request.

